# A new method to measure inter-breath intervals in infants for the assessment of apnoea and respiratory dynamics

**DOI:** 10.1101/2021.05.29.21258043

**Authors:** Tricia Adjei, Ryan Purdy, João Jorge, Eleri Adams, Miranda Buckle, Ria Evans Fry, Gabrielle Green, Chetan Patel, Richard Rogers, Rebeccah Slater, Lionel Tarassenko, Mauricio Villarroel, Caroline Hartley

**Affiliations:** Department of Paediatrics, University of Oxford, Oxford, UK; Institute of Biomedical Engineering, Department of Engineering Science, University of Oxford, Oxford, UK; Newborn Care Unit, John Radcliffe Hospital, Oxford University Hospitals NHS Foundation Trust, Oxford, UK; Department of Ophthalmology, John Radcliffe Hospital, Oxford University Hospitals NHS Foundation Trust, Oxford, UK; Department of Anaesthetics, John Radcliffe Hospital, Oxford University Hospitals NHS Foundation Trust, Oxford, UK

**Author notes:** These authors contributed equally (Ryan Purdy, João Jorge). Corresponding Author: Dr Caroline Hartley, Department of Paediatrics, University of Oxford, Level 2 Children’s Hospital, John Radcliffe, Oxford, OX3 9DU, UK.

## Abstract

**Background:** Respiratory disorders, including apnoea, are common in preterm infants due to their immature respiratory control and function compared with term-born infants. However, our inability to accurately measure respiratory rate in hospitalised infants results in unreported episodes of apnoea and an incomplete picture of respiratory dynamics.

**Methods:** We develop, validate and use a novel algorithm to identify inter-breath intervals (IBIs) and apnoeas in infants. In 42 infants (a total of 1600 hours of recordings) we assess IBIs from the chest electrical impedance pneumograph using an adaptive amplitude threshold for the detection of individual breaths. The algorithm is refined by comparing its accuracy with clinically-observed breaths and pauses in breathing. We also develop an automated classifier to differentiate periods of true central apnoea from artefactually low amplitude signal. We use this algorithm to explore its ability to identify morphine-induced respiratory depression in 15 infants. Finally, in 22 infants we use the algorithm to investigate whether retinopathy of prematurity (ROP) screening alters the IBI distribution.

**Findings:** 88% of the central apnoeas identified using our algorithm were missed in the clinical notes. As expected, morphine caused a shift in the IBI distribution towards longer IBIs, with significant differences in all IBI metrics assessed. Following ROP screening, there was a shift in the IBI distribution with a significant increase in the proportion of pauses in breathing that lasted more than 10 seconds (t-statistic=1.82, p=0.023). This was not reflected by changes in the monitor-derived respiratory rate or episodes of apnoea recorded on clinical charts.

**Interpretation:** Better measurement of infant respiratory dynamics is essential to improve care for hospitalised infants. Use of the novel IBI algorithm demonstrates that following ROP screening increased instability in respiratory dynamics can be detected in the absence of clinically-significant apnoeas.

**Funding:** Wellcome Trust and Royal Society

**Research in Context:** *Evidence before this study:* Respiratory disorders are one of the most common reasons for admission to a neonatal care unit and many pathologies and clinically-required procedures affect respiration. Despite this, current methods to measure respiratory rate in infants often provide inaccurate measurements due to factors such as poor electrode placement, movement artefact and cardiac interference. Lee and colleagues previously developed an algorithm to better identify episodes of apnoea in infants from the electrical impedance pneumograph following removal of cardiac-frequency interference. This algorithm substantially improves apnoea detection and demonstrates the high number of apnoeas that are missed in medical records. However, false apnoeas can be detected during periods of low amplitude signal caused by shallow breathing or poor electrode placement, and shorter inter-breath intervals (IBIs) cannot be assessed using the method proposed by Lee et al. limiting its use in assessing more subtle changes in an infant’s respiratory dynamics.

*Added value of this study:* We develop, test and use a new algorithm for the identification of IBIs from the electrical impedance pneumograph. We use an adaptive amplitude threshold for the identification of breaths and develop a classification model to remove periods of low amplitude signal falsely identified as episodes of apnoea. Using the algorithm, we demonstrate that retinopathy of prematurity (ROP) screening causes a significant increase in pauses in breathing that last more than 10 seconds. Our apnoea detection method was more sensitive than the current standard monitor-derived approach that is used to monitor respiratory rate in neonatal care units.

*Implications of all the available evidence:* To improve understanding of infant respiratory dynamics, better methods of assessment are essential. This will create a more complete clinical understanding of infant well-being, that will lead to improved treatment options for infants with respiratory disorders.

## Introduction

Immature respiratory control in premature infants results in irregular patterns of breathing, with frequent pauses in breathing of variable duration^1^. Apnoea (often defined as a pause in breathing lasting more than 20 seconds, or shorter if associated with a bradycardia or oxygen desaturation^2,3^) is a common pathology of prematurity, affecting more than 50% of preterm infants^3^. These events can be life-threatening, result in reduced tissue oxygenation^4^, and may have long-term effects including reduced cognitive ability in childhood^5,6^. Respiratory disorders are a common reason for admission to a neonatal unit^7^. An infant’s respiratory dynamics may also be affected by pathologies including sepsis^8^, pharmacological interventions including caffeine^9,10^ (administered as a treatment for apnoea of prematurity) and opioids^11^ (respiratory depressants), and painful clinically-indicated procedures such as retinopathy of prematurity (ROP) screening^12^. Despite the high prevalence of problems with respiratory control, clinical measurement of infant respiration is inadequate^13,14^. Whilst clinicians can rely on other vital-sign measurements to initiate the treatment of apnoeic episodes (for example, reductions in oxygen saturation and heart rate occur during prolonged pauses in breathing), self-resolving apnoeas may be missed^14^ and more subtle changes in respiratory dynamics will not be observed. Accurate assessment of respiration is essential to inform clinical practice and to understand respiratory development in health and disease.

Infants’ vital signs are continuously monitored in neonatal intensive care. Respiration is often computed by measuring changes in the electrical impedance of a patient’s thorax using the same electrodes that monitor the electrocardiograph (ECG). Commercially available vital-sign monitors use built-in algorithms to process the chest electrical impedance signal and calculate the respiratory rate, often through the identification of peaks in the signal classified as breaths as a result of a specified amplitude threshold being exceeded^15–17^. However, this approach is limited due to high-frequency noise at the cardiac frequency and artefacts caused by non-respiratory related movements^13,15,16^. Moreover, the manufacturers of many vital-sign monitors warn that their methods have yet to be validated for apnoea detection in infants^15,16^. Research investigations have demonstrated the limitations of these monitors with high false-alarm rates and missed apnoeas^13,14^. Lee and colleagues previously developed an algorithm to remove cardiac-frequency noise from the electrical impedance pneumograph (IP) signal and demonstrated improved performance compared with built-in vital-sign monitor algorithms in the detection of neonatal apnoeas^13^. However, they note that low amplitude signal related to factors such as poor electrode positioning or shallow breathing can be falsely identified as apnoeas (67% of the built-in monitor alarms in their study were found to be false; their algorithm reduced this rate to 37%)^13^. Additionally, accurate assessment of inter-breath intervals (IBIs), and not just the identification of apnoeas as in the work of Lee et al., is needed to gain a better understanding of the effects of pathology and interventions on respiration. For example, the assessment of more subtle changes in IBIs will improve classification of underlying pathology and may allow for the early detection and prediction of apnoeas^18^.

Here we develop a new method for identifying IBIs and apnoeas (defined here as pauses in breathing of at least 20 seconds) from an infant’s IP signal. We then use the algorithm to explore its sensitivity to detect changes in IBIs following morphine administration. Finally, we investigate changes in IBIs following ROP screening.

## Methods

### Study participants

A total of 42 infants were included in this study from three independent data sets. Data set 1 was collected as a subset of the MONITOR study^19^. It comprises 181 sequences of approximately 40 breaths each (in total 7,632 breaths), recorded from 5 preterm infants (post-menstrual age [PMA] at study range 30.6 – 34.3 weeks). Each breath was manually annotated by clinical staff in real time by visual observation of the infant. Data set 2 comprised vital-sign data collected during the Poppi trial, a single-centre, masked, randomised, placebo-controlled trial which investigated whether oral morphine was an effective and safe analgesic for procedural pain in premature-born infants^11^. Vital-sign data were collected for 24 hours before and after the clinical procedure – a heel lance followed by ROP screening – in 30 infants (15 received morphine, 15 received placebo, PMA at study 34-39 weeks). Data set 3 is a previously unpublished data set of 7 infants (PMA at study 30-37 weeks) whose vital signs were recorded before and after ROP screening. Further details for all studies are given in the Supplementary Methods.

All data sets were collected at the Newborn Care Unit, John Radcliffe Hospital (Oxford University Hospitals NHS Trust, Oxford, UK). Written informed parental consent was gained. Approval was obtained from South Central Research Ethics Committee (REC) (13/SC/0597) for the MONITOR study, the Medicines and Healthcare products Regulatory Agency (MHRA) and Northampton REC (15/EM/0310) for the Poppi trial, and from South Central REC (12/SC/0447) for Data set 3. All studies conformed to the standards set by the Declaration of Helsinki.

### Vital-sign recordings

All infants were monitored using a Philips IntelliVue MX800 monitor, and vital signs were continuously downloaded from the monitor using ixTrend software (ixitos GmbH, Germany). Further details are given in the Supplementary Methods.

### Breath detection from the IP signal

The algorithm presented here to identify IBIs from the IP signal consists of three main steps (Figure 1):

**Figure 1:**
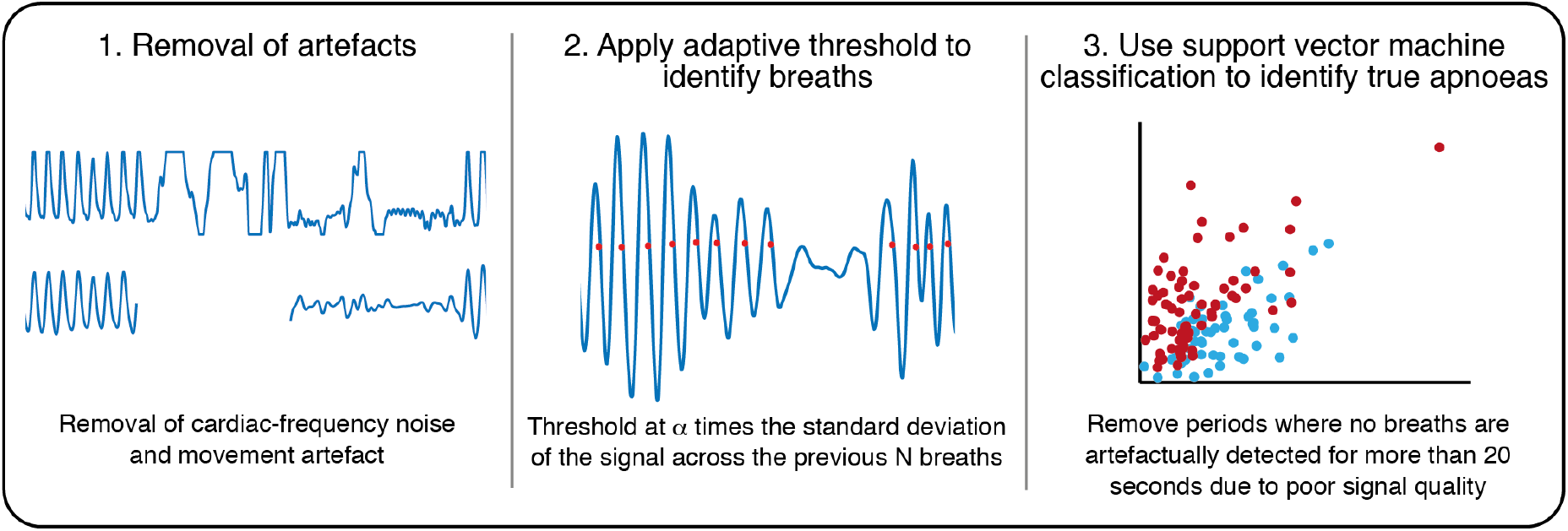
Schematic of the proposed algorithm for detection of inter-breath intervals (IBIs) from the impedance pneumograph (IP) in infants.

1. removal of artefacts,
2. application of an adaptive threshold to identify breaths,
3. identification of true apnoeas using support vector machine classification.

The code for this algorithm is available from https://gitlab.com/paediatric_neuroimaging/identify_ibi_from_ip.git. For further details of all parts of the algorithm see the Supplementary Methods, Supplementary Figures 1 and 2, and Supplementary Tables 1 and 2. Briefly, firstly, the IP signals were filtered to remove artefacts not related to respiration, for example large-amplitude changes caused by movements of the infant, and cardiac-frequency noise. Secondly, individual breaths were identified from the IP signal as the point at which an adaptive threshold is crossed (an adaptive threshold, i.e. one that is updated across the recording^20–22^, was used to account for changes in the amplitude of the signal for a variety of physiological and non-physiological reasons, such as shallow breathing and changes in the electrode and infant positioning). We identified the optimal threshold parameters for breath detection by comparing the breaths detected by the algorithm for different parameters with recordings where individual breaths were annotated in real time by a clinical member of staff visually observing the infant’s breathing (Data set 1). The optimal parameters were chosen to be values which achieved the best compromise between the percentages of false positives and false negatives. We then verified that these parameters were also suitable for detection of pauses in breathing with a duration greater than 5 seconds by comparison of pauses in breathing detected by the algorithm with those that were retrospectively identified by two investigators (Data set 2, first hour of recording, in 15 infants).

Finally, a linear support vector machine (SVM) classifier was used to identify true central apnoeas (defined here as IBIs≥ 20 seconds) as opposed to artefactually low amplitude signal. The model input features are the magnitude (root-mean-square) of the filtered IP signal during the apnoea, in the 10 seconds prior to the apnoea, and in the 10 seconds after the end of apnoea, and the change in heart rate and oxygen saturation in the 60 seconds from the onset of the apnoea. The model was trained and tested using labels (true apnoea/false alarm) provided by two investigators for all potential apnoeas identified in Data set 2 (training set, 15 infants who received morphine, test set, 15 infants who received placebo). 24% of potential apnoeas were classed differently by the two investigators and so were not included in the analysis.

### Performance of apnoea identification

To compare the accuracy of our approach with the accuracy of the current standard, all periods where the monitor-derived respiratory rate reached 0 were viewed by two investigators (see Supplementary Material) and rated according to whether the investigator thought this period was a true central apnoea or a false alarm (90% inter-rater agreement occurred here). The time of apnoeas identified by our algorithm was also compared with apnoeas documented in each infant’s clinical notes; apnoeas are documented if the clinical/nursing staff observe the infant having an apnoea.

### Use of the algorithm to evaluate respiratory depression following morphine administration

We tested the apnoea detection algorithm by examining the changes in the IBI distribution following morphine administration in the 15 infants in Data set 2 who received morphine. We examined the IBI distribution in the 1-hour period prior to drug administration with the 1-hour period after the clinical procedure (from the end of the ROP screening, on average 1.3-2.3 hours after drug administration), and calculated the mean, median, and standard deviation of the IBI distributions, the proportion of IBIs longer than 5 seconds, and the proportion of IBIs longer than 10 seconds (time periods commonly used to assess pauses in breathing^2^). We compared this with the mean monitor-derived respiratory rate calculated for the same periods.

### Use of the algorithm to evaluate changes in IBIs following ROP screening

We used the algorithm to investigate changes in the IBI distribution following ROP screening in a total of 22 infants – the 15 infants who received placebo in Data set 2 and the 7 infants in Data set 3. In the placebo-treated infants we compared the 1-hour period prior to placebo administration with the 1-hour period after the clinical procedure. In Data set 3 we similarly compared the 1-hour after ROP screening with the 1-hour period 2.3-1.3 hours prior to ROP screening. We also compared the 12-hour period before and after ROP screening in the subset of 19 infants with at least 12 hours of recording before and after ROP screening.

### Statistical analysis

All data analysis was undertaken with MATLAB 2019b (MathWorks Inc. USA). Model performance of the SVM classification was assessed with accuracy and Matthew’s correlation coefficient (MCC) using leave-one-subject-out cross-validation in the training set and independently in the test set using the model constructed from all infants in the training set. Differences in the IBI distribution and mean respiratory rate estimated from the patient monitor before and after morphine administration and ROP screening were compared using paired non-parametric t-tests with statistical significance assessed using permutation testing (10,000 random permutations) performed using FSLs PALM software^23^. P-values were adjusted for multiple comparisons using Hochberg’s method in R (The R Project for Statistical Computing).

## Results

### Optimising the adaptive threshold

We use an adaptive amplitude threshold to identify individual breaths in the IP signal. To optimise the threshold parameters, we investigated the performance of different threshold values (defined as a multiple (*α*) of the standard deviation of the IP signal across the previous N breaths) to identify individual breaths which had been visually identified at the time of recording (Data set 1). A threshold of 0.4 times the standard deviation of the filtered IP signal for the 15 previous breaths provided a good compromise between the false positive and false negative rates (Figure 2A, Supplementary Figure 3). At this threshold, a mean (across all recordings in Data set 1) of 12% of the manually-annotated breaths were missed by the algorithm (false negatives), and 13% of breaths detected by the algorithm were false positives.

**Figure 2:**
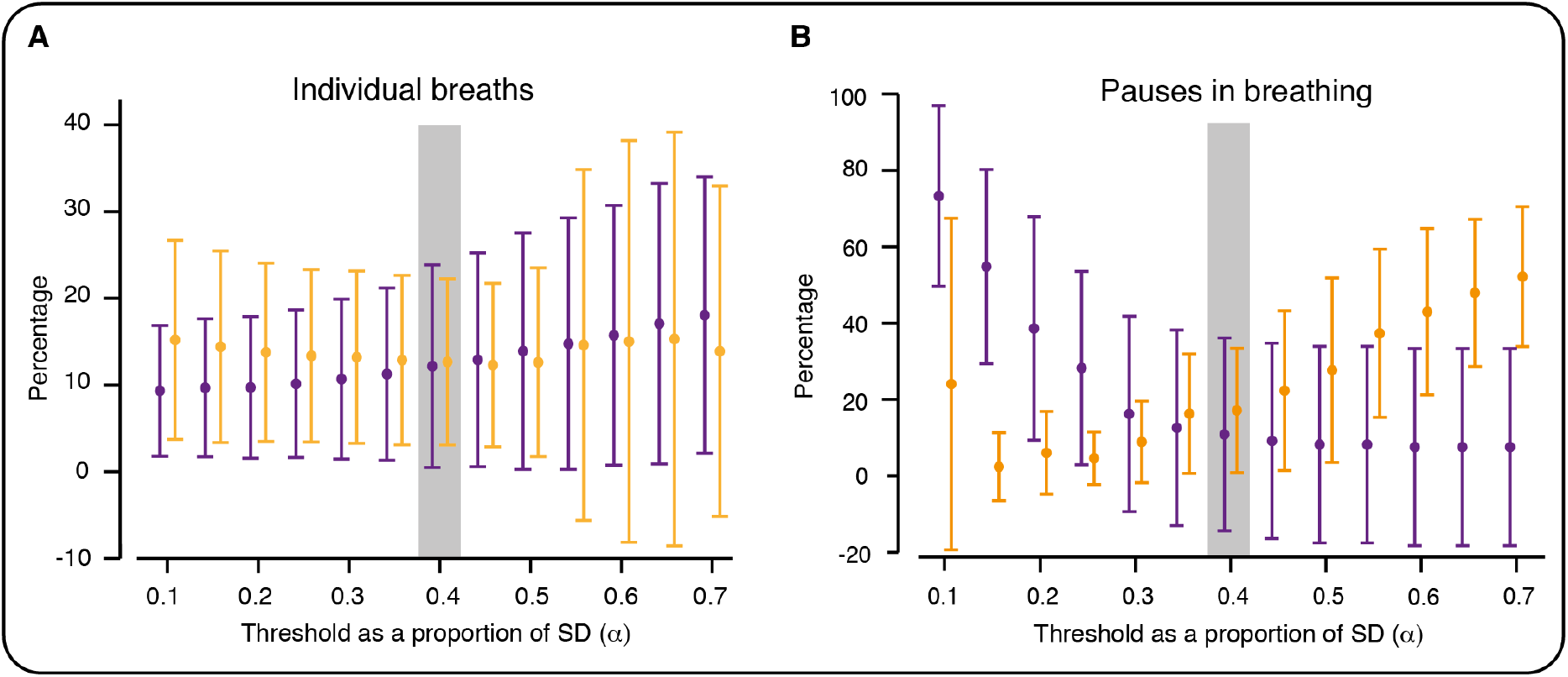
Optimising the threshold for breath detection. Percentage of false positives (orange) and false negatives (purple) for different values of α (with N=15). (A) Values calculated by comparing algorithm-identified breaths with breaths manually annotated at the time of the recording by visual observation (Data set 1). (B) Values calculated by comparing algorithm-identified pauses in breathing with pauses (of at least 5 seconds) manually annotated by two investigators (first hour of recording in 15 infants from Data set 2). Error bars indicate mean and standard deviation (across the recordings). Values are jittered on the x-axis so that false positive and false negative bars do not overlap. Grey shading indicates selected threshold parameters; with these parameters (α = 0.4, N=15), there was the optimal balance between the percentages of false positives and false negatives in the identification of individual breaths (A). These parameters also achieved a good balance between false positives and negatives in the identification of pauses in breathing (B).

We next verified whether these threshold parameters could also accurately identify pauses in breathing of at least 5 seconds (using 15 infants from Data set 2, first hour of recordings). Using the same parameters, 13 pauses out of the 162 identified by both investigators were missed by our algorithm (false negative rate: 8%) and 44 pauses out of the 229 identified by the algorithm were not identified by either investigator (false positive rate: 19%). Varying the parameters confirmed that those selected achieved a good balance between false positives and false negatives (Figure 2B).

### Optimising apnoea detection using machine learning

Shallow breathing or poor electrode placement can lead to a low-amplitude IP signal which is erroneously identified as a pause in breathing^13^. Applying the adaptive threshold to all recordings from Data set 2 identified a total of 164 potential apnoeas. Of the 164 episodes, 68 (41%) were classifed by both investigators as true apnoeas and 57 (35%) were classified by both investigators as false alarms (no agreement for 39 (24%) episodes). This already represents a major improvement in detection rate from the respiratory rate signal analysis of the patient monitor – of the 71 occasions for which the monitor-derived respiratory rate reached a value of 0 breaths per minute, two episodes were classified by both investigators as true apnoeas (3%) and 62 (87%) were classified by both investigators as false alarms.

An SVM classifier was trained to distinguish between episodes detected by the adaptive threshold and classify them as either true apnoeas or false alarms (examples shown in Figure 3A,B). In the training set (15 infants), using features derived from the IP signal alone (Figure 3C, Supplementary Methods), the classifier had an accuracy of 75% in the detection of true apnoeas versus false alarms (MCC=0.49, 62% of 69 episodes in the training set were true apnoeas). Including additional features related to the change in oxygen saturation and heart rate as inputs to the classifier and re-training it on the same training set increased the accuracy to 87% (MCC=0.74, Figure 3D).

**Figure 3:**
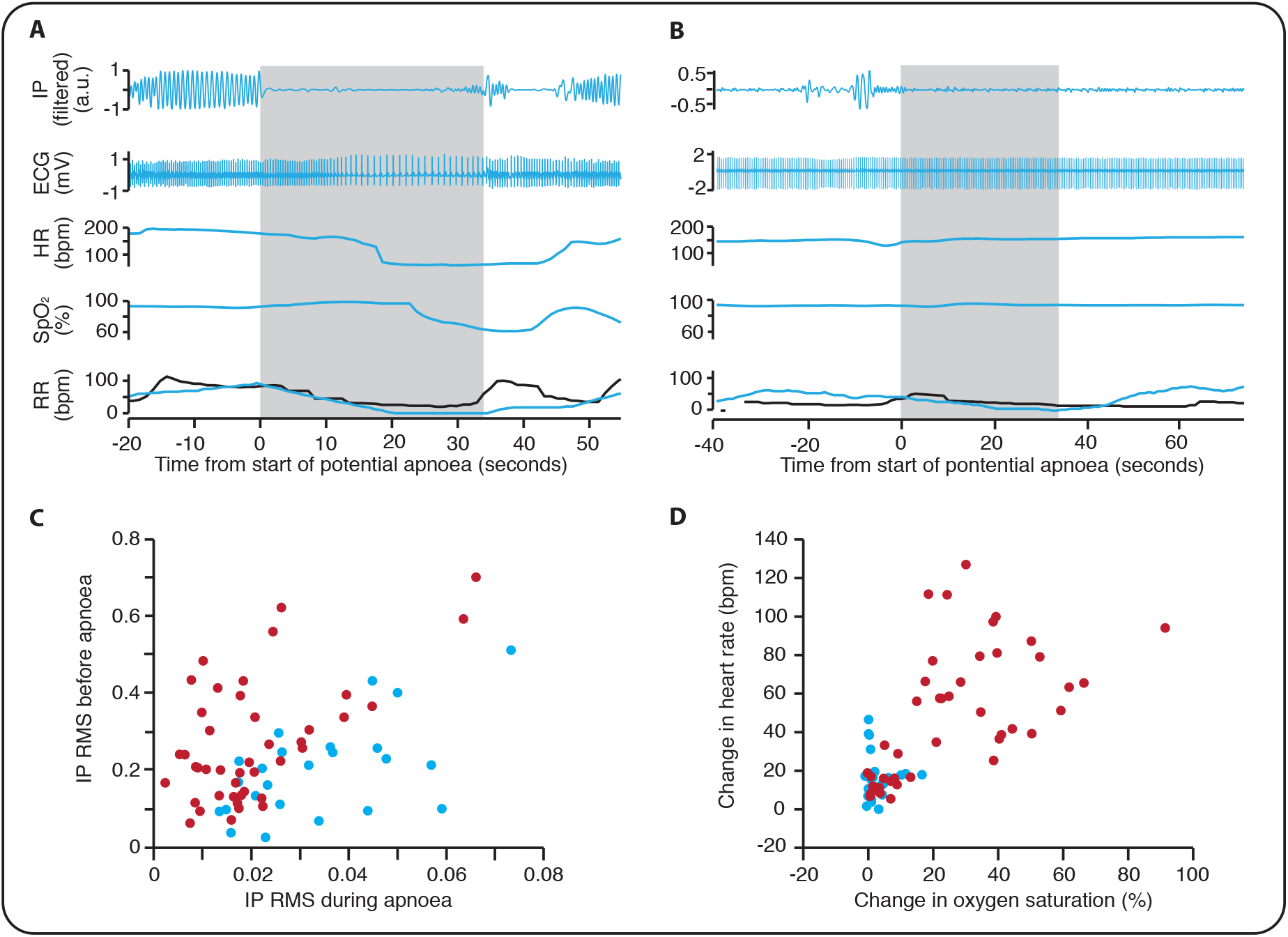
Using support vector machine classification to identify true apnoeas. (A) An example of a pause in breathing lasting longer than 20 seconds identified as a true apnoea. IP – the electrical impedance pneumograph after filtering to remove cardiac-frequency noise and movement artefact. HR – heart rate in beats per minute. SpO_2_ – oxygen saturation. RR – respiratory rate in breaths per minute, recorded by the infant’s patient monitor (black) and calculated using our algorithm (blue). Note that the RR does not reach zero on the infant’s patient monitor and so this episode does not lead to a monitor apnoea alarm. Grey shading indicates the period during which no breaths were detected by our algorithm. (B) A potential apnoea initially detected by the algorithm but classified by investigators as a false alarm. (C) The root mean square (RMS) of the IP signal before and during the apnoea (see Methods for further details). Red circles indicate episodes classified by both investigators as true apnoeas, and blue circles are those episodes classified by both investigators as false alarms. (D) Change in oxygen saturation and heart rate for true apnoeas (red) compared with false alarms (blue).

Applying the best classification model to the test set gave an accuracy of 93% (MCC=0.87, 25 of 56 episodes in the test set were true apnoeas), validating the model in this independent data set. Overall, of the 60 true apnoeas identified by our method (in both the training and test sets), 88% were missed in the clinical notes demonstrating the potential of our algorithm for improving apnoea detection. The majority of the 24 apnoeas recorded in the clinical notes were associated with an IBI of at least 10 seconds; however, 4 events were not associated with a prolonged pause in breathing but instead with a prolonged loss of signal due to artefacts. We hypothesise that such artefacts were caused by clinical intervention in response to the apnoea.

### Use of the algorithm to evaluate respiratory depression following morphine administration

Opioids are a known respiratory depressant. We used data from the morphine-treated infants in Data set 2 (from the Poppi clinical trial^11^) to check that our algorithm could identify the resulting change in IBIs. In the Poppi trial, we previously demonstrated a significant decrease in the respiratory rate (recorded on the monitor) in the morphine-treated infants compared with the placebo-treated infants, with a peak decrease approximately 2.5 hours following drug administration^11^. Thus, to compare the change in respiratory rate with changes in the IBI distribution we compared the 1-hour period prior to morphine administration with the 1-hour period following the clinical procedure (approximately 1.3-2.3 hours after drug administration). As expected, there was a significant decrease in the respiratory rate recorded by the patient monitor (*p*=0.0004, non-parametric permutation t-test corrected for multiple comparisons, n=15, Table 1, Figure 4A). This was reflected in the IBI distribution, which showed a clear shift in the distribution towards longer IBIs following the clinical procedure (Figure 4B), and significant differences in all IBI metrics assessed (Figure 4C, D, Table 1).

**Table 1:** Changes in inter-breath intervals following morphine administration and ROP screening. Comparison of the respiratory rate (recorded by the patient monitor) and inter-breath interval (IBI) distribution one hour before and after morphine administration and one hour before and after ROP screening. The table indicates the mean across all infants in each group, and the t-statistic and p-values for each comparison (permutation test). *P*-values were corrected for multiple comparisons using Hochberg’s method (* indicates corrected *p*<0.05, ** *p*<0.01, *** *p*<0.001).

|  | Mean<br>Before | Mean<br>After | t-statistic | Uncorrected<br>p-value | Corrected<br>p-value |
| --- | --- | --- | --- | --- | --- |
| **Morphine (n=15 infants)** |  |  |  |  |  |
| Mean respiratory rate (bpm) | 52.01 | 44.66 | -3.54 | 0.0001 | 0.0004 *** |
| Mean IBI (seconds) | 1.07 | 1.34 | 5.18 | 0.0001 | 0.0004 *** |
| Median IBI (seconds) | 0.93 | 1.03 | 3.96 | 0.0012 | 0.0012 ** |
| Standard deviation IBI (seconds) | 0.61 | 1.26 | 5.86 | 0.0001 | 0.0004 *** |
| IBI > 5 seconds (%) | 0.56 | 2.54 | 4.17 | 0.0004 | 0.0008 *** |
| IBI > 10 seconds (%) | 0.02 | 0.49 | 3.39 | 0.0002 | 0.0006 *** |
| **ROP screening (n=22 infants)** |  |  |  |  |  |
| Mean respiratory rate (bpm) | 51.07 | 50.92 | -0.14 | 0.89 | 0.89 |
| Mean IBI (seconds) | 1.09 | 1.12 | 1.32 | 0.20 | 0.81 |
| Median IBI (seconds) | 0.97 | 0.98 | 0.25 | 0.84 | 0.89 |
| Standard deviation IBI (seconds) | 0.56 | 0.66 | 2.45 | 0.021 | 0.10 |
| IBI > 5 seconds (%) | 0.49 | 0.63 | 1.14 | 0.28 | 0.83 |
| IBI > 10 seconds (%) | 0.02 | 0.06 | 1.82 | 0.0039 | 0.023 * |

**Figure 4:**
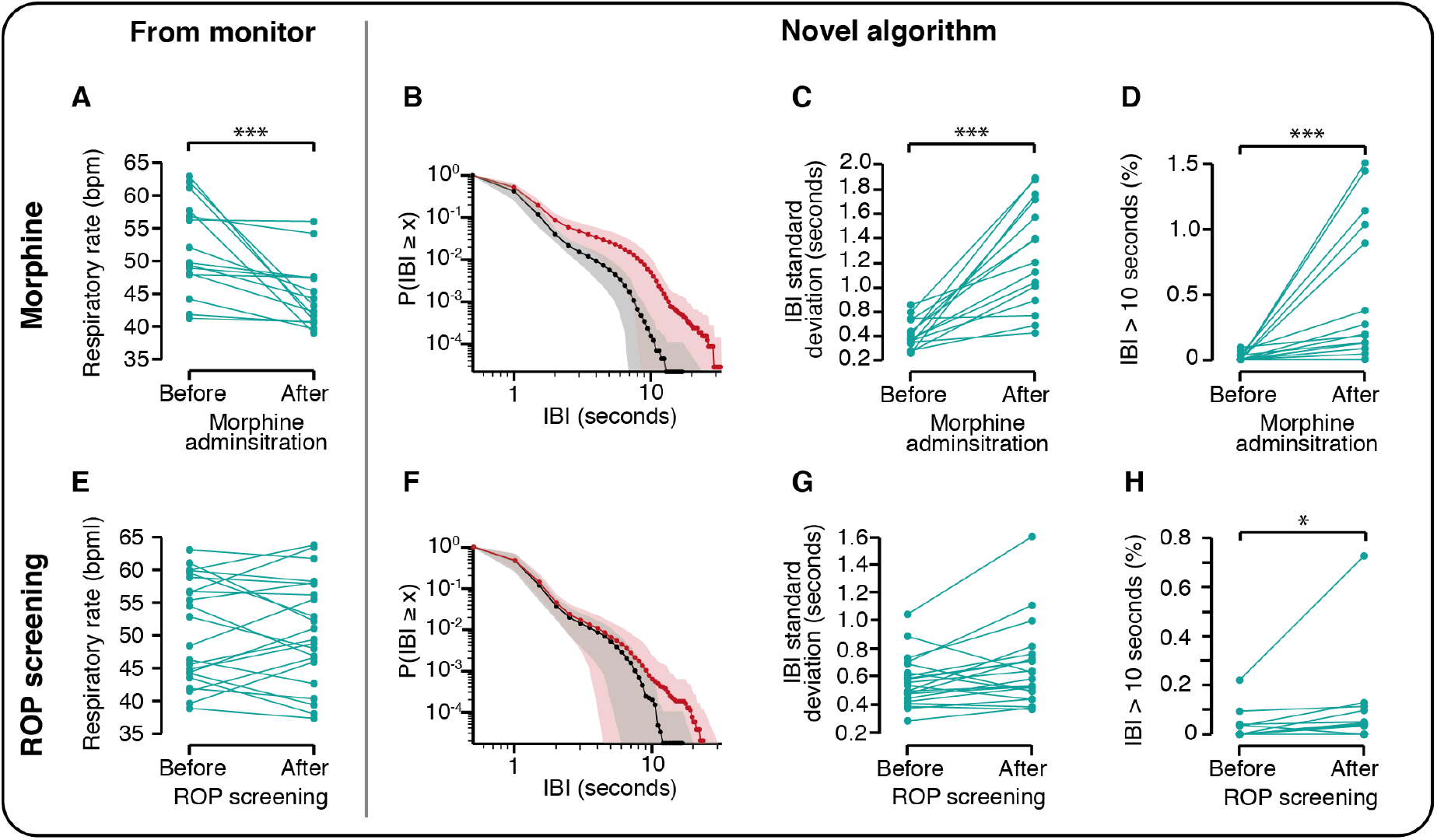
Inter-breath intervals are altered by morphine administration and following ROP screening. (A-D) Respiratory rate and inter-breath intervals (IBIs) in the one-hour period prior to morphine administration compared with a one-hour period after morphine administration (the one-hour period immediately following ROP screening, approximately 1.3-2.3 hours after morphine administration) in the 15 infants who received morphine in the Poppi clinical trial. (E-H) Respiratory rate and IBIs one hour before and after ROP screening in 22 infants. (A, E) Mean respiratory rate from the infants’ patient monitor. (B-D, F-H) Metrics calculated using the novel algorithm proposed in this paper to identify the IBIs. (B) IBI distribution in the one-hour period prior to (black) compared with 1.3 – 2.3 hours after morphine administration (red). (F) IBI distribution in the one-hour period before (black) and after (red) ROP screening. Y-axis indicates the probability of an IBI of duration greater than or equal to the x-axis value. Dotted line indicates the mean and shaded area the standard deviation. (* indicates *p*<0.05, ** *p*<0.01, *** *p*<0.001, *p*-values corrected for multiple comparisons).

### Use of the algorithm to evaluate changes in IBIs following ROP screening

ROP screening, an eye exam that is thought to be painful and distressing for infants^24^, has previously been shown to increase the rate of apnoea in the 24 to 48 hours following the screen^12^. Using our algorithm, we compared the IBI distribution one-hour before and after ROP screening in 22 infants. Infant demographics are shown in Table 2. There was a shift in the IBI distribution in the 1-hour following ROP screening towards longer IBIs (Figure 4F), with a significant increase in the proportion of IBIs longer than 10 seconds (*p*=0.023, Figure 4H, Table 1). This was not reflected by a change in the respiratory rate recorded by the patient monitor (*p*=0.89, Figure 4E, Table 1). Moreover, in the subset of 19 infants who had vital signs recorded for at least 12 hours before and after ROP screening, there was a significant increase in the proportion of IBIs longer than 10 seconds in the 12 hours after ROP screening compared with the 12 hours before (*p*=0.037, t-statistic=1.77). No apnoeas were recorded in the clinical notes in the 12 hours before or after ROP screening for any of these infants.

**Table 2:** Demographic details for the 22 infants where inter-breath interval distributions are compared before and after ROP screening. IQR – interquartile range, SD – standard deviation, * Apgar scores were missing from the notes of 2 infants.

|  |  |
| --- | --- |
| Gestational age at birth (weeks) – median (IQR) | 28.1 (27.1 – 29.3) |
| Postmenstrual age at study (weeks) – median (IQR) | 34.9 (34.3 – 36.1) |
| Weight at birth (g) – median (IQR) | 1100 (889 – 1228) |
| Weight at study (g) – median (IQR) | 2080 (1923 – 2206) |
| Sex – Female/Male – number (%) | 12 (55) / 10 (45) |
| Normal vaginal delivery – number (%) | 7 (32) |
| Assisted delivery – number (%) | 1 (4) |
| Caesarean delivery – number (%) | 14 (64) |
| Apgar score at 10 minutes – mean (SD)* | 8.9 (1.5) |
| Infants ventilated during admission – number (%) | 15 (68%) |
| Number of days ventilated in those ventilated – median (IQR) | 2 (1.5-11.5) |

## Discussion

We developed a new algorithm to detect IBIs from the IP signal in infants. We used an adaptive amplitude threshold to identify individual breaths, validating the threshold by comparison with visually identified breaths and pauses in breathing. Previous studies have reported that signals with low amplitude due to poor electrode placement or shallow breathing can be erroneously detected as episodes of apnoea. To overcome this problem, we used machine learning to identify true apnoeas from periods of artifactually low amplitude. We tested our algorithm by investigating changes in IBIs following morphine administration, observing a clear shift in the IBI distribution consistent with the reduction in respiratory rate seen on the infants’ patient monitors. Finally, we used our algorithm to investigate changes in IBIs following ROP screening. We observed a significant shift in the IBI distribution following ROP screening which was not reflected by a change in the respiratory rate recorded by the monitors. This demonstrates the increased sensitivity of our method and highlights the increase in physiological instability in infants following ROP screening.

Premature infants are born with immature cerebral and respiratory function compared with term-born infants, and consequently have a higher incidence of respiratory disorders. Current inadequacies in the measurement of respiration in infants leads to missed opportunities to better understand respiratory development and could potentially lead to suboptimal clinical treatment. For example, caffeine therapy, given for apnoea of prematurity, is stopped in infants between 33-35 weeks PMA if the infant appears clinically stable^25^. However, in 10% of infants caffeine treatment is restarted^26^, which may suggest that caffeine was withdrawn too early, exposing the infants to the adverse consequences of lack of treatment. We found that 88% of apnoeas identified using our algorithm were missed in the clinical notes. Improved measurement of respiration is essential to optimise clinical treatment of apnoea and could enhance treatment for other clinical conditions or procedures which alter respiration.

Many drugs will alter infants’ vital signs. Our results confirm the applicability of the algorithm to analyse morphine-related respiration depression. Using this approach to investigate respiratory changes in relation to other drugs commonly prescribed in neonatal care may enhance our understanding of pharmacodynamics. Additionally, analysis of vital signs may be useful to develop predictive models to tailor individualised care^27,28^. We recently showed in a post-hoc analysis of the morphine-treated infants in the Poppi trial that we could predict the risk of adverse cardiorespiratory effects in individual infants from their baseline physiological stability^29^. To date, measures of respiration are often not included in the development of predictive tools, which is likely due to the relatively poor quality of the currently-available measurement tools^28^. Here we provide a more accurate measure of IBIs, which will allow for more complex metrics, such as respiratory rate variability, to be computed.

ROP screening is a routine procedure that is performed approximately every 2 weeks in infants born very prematurely. Previous research has suggested that there is an increase in apnoeas following screening from clinical chart review^12^. In exploratory analysis, we demonstrated a significant increase in the proportion of IBIs longer than 10 seconds in the 1-hour and 12-hour periods after ROP screening, which was not reflected by a change in the respiratory rate recorded on the monitor or by apnoeas recorded on clinical charts. This demonstrates the improved sensitivity of our method for identifying changes in respiratory dynamics and suggests that even those infants without clinically-significant apnoeas may still experience changes in respiratory dynamics with a shift towards longer IBIs. Further research in a larger cohort across a wider age range is needed to explore the relationship between an infant’s respiratory dynamics following ROP screening and changes with age. Identifying older infants that are at risk of physiological instability after ROP screening would be particularly important for those ex-premature infants who have ROP screening in outpatient clinics and may benefit from observation before leaving hospital^30^.

By using an adaptive threshold which we validated for infants, and combining this with machine learning classification, our algorithm performed substantially better than the monitor derived respiratory rate in identifying apnoeas. However, limitations of this study are the relatively small sample size and narrow age range of the infants included (from 30-39 weeks PMA). Further validation should be carried out in younger infants. Moreover, this method identifies central apnoea; it cannot detect obstructive apnoea – alternative measures, such as nasal air flow, are needed to detect these events. Additionally, apnoea that necessitates intervention by clinical staff may not be detected or the reported duration may be shorter than the true duration of the episode as interventions are likely to lead to large artefacts in the IP signal. Whilst this is not a problem for clinical management, as the infant is receiving the appropriate clinical intervention to support their breathing, this should be taken into account in research studies so that apnoeas are not missed in the analysis.

In summary, despite the common occurrence of respiratory pathology in preterm infants, current methods used to measure respiration are inadequate. We developed a new method to measure respiration in infants, demonstrating the improved sensitivity of the method compared with current standards. Furthermore, we identified a significant increase in respiratory instability in infants following ROP screening. A better understanding of respiratory dynamics in infants is critical to improve neonatal care.

## Data Availability

All data reported in this manuscript is available upon request from the corresponding author.

https://gitlab.com/paediatric_neuroimaging/identify_ibi_from_ip.git.

## Author contributions

CH conceived the idea for the study. TA, RP, JJ, REF, RR, MV and CH conducted the analysis. TA, RP, JJ, RR, RS, MV and CH interpreted the data. TA and CH wrote the first draft of the manuscript. Data set 1 was originally collected for the MONITOR study by JJ, GG, MV, LT and members of the MONITOR study team. Data set 2 was originally collected for the Poppi Trial by CH, GG, MB, RR, CP, EA, RS and members of the Poppi trial team. Data set 3 was collected by MB and CP. All authors critically reviewed the data and revised the paper.

## Acknowledgements

This work was funded by the Wellcome Trust and Royal Society through a Sir Henry Dale Fellowship (grant reference number: 213486/Z/18/Z). We would like to thank the MONITOR Trial team for collecting Data set 1, conducting analysis and interpreting results as part of the original MONITOR study. We would like to thank the Poppi Trial team for collecting Data set 2, conducting analysis and interpreting results as part of the original Poppi trial. We would also like to thank Gabriela Schmidt Mellado for assistance with data collection for Data set 3, Fahiza Begum for assistance with analysis in an early form of this algorithm, and Prof. John Delos for sharing the code related to Lee et al. *A new algorithm for detecting central apnea in neonates. Physiol Meas* 2012; 33: 1–17.

## References

1 Martin RJ, Abu-Shaweesh JM. Control of breathing and neonatal apnea. In: Biology of the Neonate. 2005. DOI:10.1159/000084876.

2 Elder DE, Campbell AJ, Galletly D. Current definitions for neonatal apnoea: Are they evidence based? J Paediatr Child Health 2013; 49: E388–96.

3 Finer NN, Higgins R, Kattwinkel J, Martin RJ. Summary proceedings from the apnea-of-prematurity group. Pediatrics 2006; 117: S47–51.

4 Yamamoto A, Yokoyama N, Yonetani M, Uetani Y, Nakamura H, Nakao H. Evaluation of change of cerebral circulation by SpO2 in preterm infants with apneic episodes using near infrared spectroscopy. Pediatr Int 2003; 45: 661–4.

5 Janvier A, Khairy M, Kokkotis A, Cormier C, Messmer D, Barrington KJ. Apnea is associated with neurodevelopmental impairment in very low birth weight infants. J Perinatol 2004; 24: 763–8.

6 Pillekamp F, Hermann C, Keller T, Von Gontard A, Kribs A, Roth B. Factors influencing apnea and bradycardia of prematurity - Implications for neurodevelopment. Neonatology 2007. DOI:10.1159/000097446.

7 Gallacher DJ, Hart K, Kotecha S. Common respiratory conditions of the newborn. Breathe 2016; 12: 30–42.

8 Fairchild K, Mohr M, Paget-Brown A, et al. Clinical associations of immature breathing in preterm infants: Part 1-central apnea. Pediatr Res 2016; 80: 21–7.

9 Moschino L, Zivanovic S, Hartley C, Trevisanuto D, Baraldi E, Roehr CC. Caffeine in preterm infants: where are we in 2020? ERJ Open Res 2020; 6: 00330–2019.

10 Schmidt B, Roberts RS, Davis P, et al. Caffeine therapy for apnea of prematurity. N Engl J Med 2006; 354: 2112–21.

11 Hartley C, Moultrie F, Hoskin A, et al. Analgesic efficacy and safety of morphine in the Procedural Pain in Premature Infants (Poppi) study: randomised placebo-controlled trial. Lancet 2018; 392: 2595–605.

12 Mitchell AJ, Green A, Jeffs DA, Roberson PK. Physiologic effects of retinopathy of prematurity screening examinations. Adv Neonatal Care 2011; 11: 291–7.

13 Lee H, Rusin CG, Lake DE, et al. A new algorithm for detecting central apnea in neonates. Physiol Meas 2012; 33: 1–17.

14 Vergales BD, Paget-Brown AO, Lee H, et al. Accurate automated apnea analysis in preterm infants. Am J Perinatol 2014; 31: 157–62.

15 Koninklijke Philips. IntelliVue Patient Monitor. 2019. https://www.fda.gov/media/137229/download (accessed Oct 8, 2020).

16 Draeger. Infinity Acute Care System: Instructions for use. 2017. https://www.draeger.com/Products/Content/iacs-vg7-monitoring-applications-ifu-ms34093-en.pdf (accessed Oct 8, 2020).

17 Welch Allyn 1500 Patient Monitor. 2013. https://www.welchallyn.com/content/dam/welchallyn/documents/upload-docs/Training-and-Use/User-Manual/Welch-Allyn-1500-Patient-Monitor-Software-version-1.4.X_User-Manual.pdf (accessed Oct 8, 2020).

18 Lim K, Jiang H, Marshall AP, Salmon B, Gale TJ, Dargaville PA. Predicting Apnoeic Events in Preterm Infants. Front Pediatr 2020; 8: 1–7.

19 Jorge J, Villarroel M, Chaichulee S, Green G, McCormick K, Tarassenko L. Assessment of signal processing methods for measuring the respiratory rate in the neonatal intensive care unit. IEEE J Biomed Heal Informatics 2019; 23: 2335–46.

20 Zong W, Moody GB, Jiang D. A robust open-source algorithm to detect onset and duration of QRS complexes. In: Computers in Cardiology. 2003: 737–40.

21 Zong W, Heldt T, Moody GB, Mark RG. An open-source algorithm to detect onset of arterial blood pressure pulses. In: Computers in Cardiology. 2003: 259–62.

22 Karlen W, Ansermino JM, Dumont G. Adaptive pulse segmentation and artifact detection in photoplethysmography for mobile applications. In: Proceedings of the Annual International Conference of the IEEE Engineering in Medicine and Biology Society, EMBS. 2012: 3131–4.

23 Winkler AM, Ridgway GR, Webster MA, Smith SM, Nichols TE. Permutation inference for the general linear model. Neuroimage 2014; 92: 381–97.

24 Belda S, Pallás CR, De La Cruz J, Tejada P. Screening for retinopathy of prematurity: Is it painful? Biol Neonate 2004; 86: 195–200.

25 National Institute for Health and Care Excellence. Specialist neonatal respiratory care for babies born preterm. NICE Guideline NG124. 2019.

26 Haddad W, Sajous C, Hummel P, Guo R. Discontinuing caffeine in preterm infants at 33-35 weeks corrected gestational age: Failure rate and predictive factors. J Neonatal Perinatal Med 2015; 8: 41–5.

27 Griffin MP, O’Shea TM, Bissonette EA, Harrell FE, Lake DE, Moorman JR. Abnormal heart rate characteristics preceding neonatal sepsis and sepsis-like illness. Pediatr Res 2003; 53: 920–6.

28 Kumar N, Akangire G, Sullivan B, Fairchild K, Sampath V. Continuous vital sign analysis for predicting and preventing neonatal diseases in the twenty-first century: big data to the forefront. Pediatr Res 2020; 87: 210–20.

29 Hartley C, Baxter L, Moultrie F, et al. Predicting severity of adverse cardiorespiratory effects of morphine in premature infants: a post hoc analysis of Procedural Pain in Premature Infants trial data. Br J Anaesth 2020; S0007-0912: 30913–2.

30 Wood MG, Kaufman LM. Apnea and bradycardia in two premature infants during routine outpatient retinopathy of prematurity screening. J AAPOS 2009; 13: 501–3.

