## Supplementary Material for "A new method to measure inter-breath intervals in infants for the assessment of apnoea and respiratory dynamics"

### Supplementary Methods

*Study design and participants*

Full details of data collection for the MONITOR study (Data set 1) are given in Jorge et al. 2019[1]. This data was collected in five infants aged between 30 – 34 weeks postmenstrual age (PMA) at the time of study. Each infant's vital signs were recorded for between 2-4 days, during which time manual counts of the infants breathing were recorded. Manual counts consisted of a train of approximately 40 breaths which were manually annotated onto the recording by a clinical member of staff who watched the infant and pressed the space key on the study computer each time they observed a full breath. A total of 181 manual count trains from the five infants were assessed in the present study.

Full details of the recruitment, trial design and trial procedures for Data set 2 from the Poppi (Procedural Pain in Premature Infants) trial are given in Hartley and Moultrie et al. 2018[2–4]. Briefly, infants were aged between 34-39 weeks PMA at the time of study and had been born at less than 32 weeks' gestation or with a birth weight lower than 1501 g, fulfilling national criteria for retinopathy of prematurity (ROP) screening. Infants were given either oral morphine (100 µg/kg, n=15 infants) or a placebo (n=15) approximately 1 hour prior to the clinical procedure - a medically-required heel lance for blood sampling and a ROP screening. As part of the trial, vital-sign data was recorded for approximately 24 hours before and after the clinical procedure (total duration of recordings: 1503 hours). The trial was stopped early due to profound adverse respiratory effects of morphine in the study participants (requiring treatment with non-invasive positive pressure ventilation).

Data set 3 comprised of data from 7 infants who were aged between 30-37 weeks PMA at the time of study and had been born at less than 32 weeks' gestation or with a birthweight lower than 1501 g, fulfilling national criteria for ROP screening. This was a subset of infants collected as part of an ongoing study to investigate changes in brain activity and vital signs following ROP screening. Infants from this study were selected for inclusion if they had a minimum of one hour of vital signs recording before and after ROP screening. All 7 infants selected had recordings for at least 12 hours before ROP screening. Two infants were recorded for approximately 1 hour after ROP screening, one infant for 9 hours and the other 4 infants at least 12 hours recording after ROP screening.

For infants in both Data set 2 and 3, ROP screening was performed using binocular indirect ophthalmoscopy. Mydriatic eye drops (tropicamide 1% and phenylephrine 2.5%) were administered approximately 60 minutes and again at approximately 45 minutes before the examination. Prior to the examination the infant was swaddled, and immediately prior topical local anaesthetic (proxymetacaine 0.5%) eye drops were instilled. For the examination an eyelid speculum was used, and binocular indirect ophthalmoscopic examination was completed using a Flynn style indenter.

*Vital-sign recordings*

Heart rate, oxygen saturation and respiratory rate (calculated by the monitor) were downloaded at a sampling rate of 1 Hz; the ECG was recorded at a sampling rate of 250 Hz from 3 electrodes placed on the infant's chest, and the IP at a sampling rate of 62.5 Hz from the chest electrodes. In Data set 2 the time of drug administration, heel lancing, and ROP screening, and in Data set 3 the time of ROP screening, were electronically marked on the physiological recordings by a researcher in real time.

*Development of a new algorithm to identify breaths from the IP signal*

The code for this algorithm is available from
https://gitlab.com/paediatric_neuroimaging/identify_ibi_from_ip.git.

Removal of noise and artefacts

Cardiac-frequency noise is a particular problem affecting IP signals as the cardiac-synchronous signal can be erroneously detected as respiration, particularly during pauses in breathing (Supplementary Figure 1A, B). To remove this artefact, we used an approach similar to Lee and colleagues[5], whereby the timing of R-peaks is first identified from the ECG signal and from this the series of the time intervals between consecutive R-peaks, known as RR intervals, is computed. Lee et al. proposed a novel approach of resampling of the IP signals according to the timings of the RR intervals. (a) The time scale is digitally stretched or contracted such that RR intervals are equally spaced, and thus the unit of time becomes one RR interval rather than 1 second. (b) Using the Fourier transform, the R-peak contaminants within the IP would then be easily identifiable in the frequency domain, at 1 Hz, and the harmonics of 1 Hz, and could easily be removed. (c) The inverse Fourier transform would then be computed, giving an IP signal with the cardiac-frequency noise removed. In practice, the three steps (a), (b) and (c) are combined into a single step in the time domain using either band-stop or notch filters.

Empirically we found that the filter regime introduced by Lee et al. did not remove the R-peak interference from the IP signals in our data. This was primarily due to the poor performance of the Pan-Tompkins algorithm[6,7], which was used by Lee et al. to identify ECG R-peaks[5], on our IP data. Instead, we used signal envelopes to identify R-peaks, whereby the envelope of the ECG signal was generated using the MATLAB *envelope* function (which uses a spline interpolation of the signal maxima), in 5-second segments, enabling the identification of the R-peaks as the points of upper intersect between the envelope and the ECG signal. The RR intervals derived from the identified R-peaks were then stretched into 50 equally-sized steps, before resampling the IP signals in accordance with the RR intervals, as described above. Two infinite impulse response (IIR) notch filters were used to remove the 1 Hz and 2 Hz components (representing the R-peak interference) from the IP signal, before resampling the signal at 50 Hz. Following this, low frequency movement artefacts were removed using a high-pass finite impulse response (FIR) filter with a cut-off frequency of 0.5 Hz. Finally, large amplitude artefacts in the ECG recording could lead to large amplitude artefacts in the IP signal when it is filtered using the ECG signal. Extreme outliers in the filtered IP signals were identified through visual assessment as signal values with amplitudes greater than 6 times the 90$^{th}$ percentile of the positive values of the filtered signal or less then 6 times the 10$^{th}$ percentile of the negative values of the filtered signal; these were replaced with these boundary values. This occurred in 0.0009% of the total signal recorded across all infants in Data set 2. The differences between the computation employed by Lee et al. and that used in this study are summarised in Supplementary Table 2.

The IP signal recorded on the monitor is hard-limited at upper and lower values. Gross movement artefact or periods of interference can lead to the signal being hard-limited for some period of time (example shown in Supplementary Figure 1C). To avoid these segments of hard-limited signal being erroneously detected as pauses in breathing in the filtered signal, segments of IP which corresponded to periods of which the original IP signal remained at a continuous maximum or minimum value for at least 1 second were removed. In addition, the 2.5-second segments either side of these segments were also removed.

Gross artefacts which affect the ECG (and so also the IP signal) are detected by the patient monitor, which will not calculate a heart rate value during this period. Therefore, segments of IP which corresponded to instances of missing estimates of HR, and the 2.5-second segments of the IP 2.5 seconds either side of these episodes, were also removed from the IP signal.

Identification of breaths

The breath waveforms were segmented from the filtered IP signal as the point at which the signal crossed an amplitude threshold, and pauses in breathing were defined as inter-breath intervals of 5 seconds or longer. The threshold, $\theta$, used to identify breaths was set as a proportion of the standard deviation of the IP signal across the previous N breaths. That is $\theta = \alpha\sigma(IP_N)$, where $IP_N$ is the IP signal for the N previous breaths, and $\alpha$ is a scaling factor. We identified optimal values of $\alpha$ and N that achieved a balance between the number of false positives and false negatives in the identification of pauses in breathing and individual breaths (see Threshold optimisation). The first 600 seconds of the IP signal were analysed using a fixed threshold of $\theta = \alpha\sigma(IP|_0^{600})$, and the adaptive threshold was used for the rest of the recording. Threshold crossings occurring within 0.3 seconds of the previous crossing were removed.

From this time series of breaths, the time intervals between consecutive breaths were computed giving the sequence of inter-breath-intervals (IBIs) for each infant. An IBI of 5 seconds or more was defined as a pause in breathing. Pauses in breathing which occurred within 2 seconds of a previous pause were combined and counted as a single pause. Also, as it is possible that a pause in breathing could begin during a period in which the IP data has been removed or is missing, IBIs of 5 seconds or more occurring immediately before or after a gap in IP data were included in the IBI series.

The time series of breaths was also used to compute the infants' respiratory rate shown in Figure 3. The respiratory rate was calculated from the number of breaths in a 20 second sliding window, moved through the IP signal with a 1-second increment; thus, a respiratory rate of 0 corresponds to an apnoea lasting 20 seconds or more.

Threshold optimisation

To determine the optimal threshold parameters for the identification of breaths in the infants' IP signals we assessed the algorithm's ability to detect individual breaths using Data set 1 for different levels of $\alpha$ and N. The manually annotated breaths were defined as true breaths, and false negatives defined as true breaths not identified by the algorithm. False positive breaths were defined as the breaths identified by the algorithm, but not annotated by clinical staff. Manual annotations were not necessarily annotated at a specific time within the breath wave and so the time of the manual annotations and the threshold crossings would not be expected to match exactly. However, the timings would be expected to be sequential i.e. there should only be one manual breath annotation between any two threshold crossings and vice versa. Therefore, the presence of two successive manual breath annotations without a threshold crossing between them indicated that the algorithm did not identify a breath (false negative), whilst two successive threshold crossings without a manual breath annotation between them indicated that the algorithm identified a false positive breath. The IP signal epochs from Data set 1 were each only approximately 180 seconds long (60 seconds before and approximately 80 seconds after a train of manual counts), so the threshold was initially applied to the first 40 seconds of data, before using an adaptive threshold. Initially, the optimal value for $\alpha$ was identified as that which achieved a balance (i.e. similar number) between the false positive and false negative rates. In this assessment $\alpha$ was varied from 0.1 to 0.7 (with increments of 0.05) and a fixed value of N=15 was used. Having identified an optimal value for $\alpha$, N was then varied to identify an optimal value for this parameter (which was identified as N=15).

To verify that the chosen threshold parameters were also able to accurately identify pauses in breathing we also compared manual ratings of pauses in breathing with pauses in breathing identified by the algorithm. Pauses in breathing of greater than 5 seconds were identified in the first hour of the recordings in 15 of the infants from Data set 2 (of the 15 infants studied, 8 received morphine and 7 received placebo, though no drug had been given during the section of the recording assessed) by two investigators experienced in reviewing neonatal vital-sign data. The investigators identified pauses in breathing through visual assessment of the infants' original IP signals directly from the monitor and the filtered IP signals (with cardiac-frequency noise and movement artefacts removed). Pauses in breathing where there were single oscillations (possibly breaths/gasps or possibly artefact from movement or stimulation) within a pause were identified as one pause. The investigators also reviewed the ECG, heart rate, and oxygen saturation traces for the infant, though the identification of pauses in breathing was primarily based on their visual inspection of the IP signals as short pauses in breathing do not necessarily affect the infant's heart rate or oxygen saturation.

For each threshold parameter, the pauses in breathing identified by the algorithm were compared to those identified by the two investigators; the pauses found by both investigators were defined as true pauses. The false negative rate was defined as the percentage of true pauses not identified by the algorithm. The false positive rate was defined as the percentage of pauses identified by the algorithm but identified by neither investigator.

Occasionally, due to technical difficulties, periods occurred where the ECG signal was not recorded but the IP signal was (0% in Data set 1, 0.3% of the total ECG record in Data set 2, 0.4% of the total record in Data set 3). Thus, during these periods, cardiac-frequency noise could not be removed from the IP signal. To assess the performance of the adaptive threshold without the removal of cardiac-frequency noise we also applied the algorithm to the same sections of the recording (which had ECG recorded throughout), for both the identification of breaths (Data set 1) and the identification of pauses in breathing (first hour of recordings in 15 infants from Data set 2) but did not filter out the cardiac noise.

Accurate identification of apnoeas using machine learning

Shallow breathing or poor electrode placement can lead to a low amplitude IP signal which is erroneously detected as a prolonged pause in breathing. We used machine learning to identify episodes of central apnoea versus periods erroneously detected as a prolonged pause in breathing. To develop the model, two investigators viewed the original IP signal, the filtered IP signal, the ECG, the heart rate and the oxygen saturation for 30 seconds before and after the start of all potential apnoeas identified in the entire recordings from all 30 infants in Data set 2. We defined here a central apnoea as a pause in breathing lasting 20 seconds or longer, so the investigators evaluated all periods for which a breathing rate of 0 was estimated. The investigators determined whether they considered each potential apnoea to be a true apnoea or a false alarm (i.e. whether there was low amplitude signal for another reason such as shallow breathing or poor electrode placement). The investigators viewed a total of 164 episodes of potential apnoea from the 30 infants and had an inter-rater agreement of 76%.

A support vector machine classification model was developed using the data from the 15 morphine-treated infants (training set), with all 69 episodes for which the investigators agreed, with 62% of these episodes classified as true apnoeas and the other 38% being classified as false alarms. Model performance was assessed using leave-one-subject-out cross-validation (i.e. all episodes for a subject were removed in a single fold). Having trained the model on the data from the morphine-treated infants, it was then tested in the data from the 15 placebo-treated infants (test set).

The model was constructed with 5 features:

1. The root-mean-square (RMS) of the IP signal during the episode (possible apnoea). The RMS was calculated in 1-second intervals from 1 second after the last annotated breath, until 19 seconds after the last annotated breath (i.e. up until the point immediately before the pause in breathing is classed as an apnoea according to our definition of at least 20 seconds in length), and the median of the RMS values across all intervals selected. The first and last second were removed as the IP signal can be affected by the edge of the last/first breaths as the time of first/last breath is selected as the point at which the threshold is crossed (Supplementary Figure 2A). The median value from the RMS values of the filtered IP signal was computed for each 1-second interval, as movement artefact in the IP signal could increase a sub-set of the RMS values even if a real apnoea was occurring (Supplementary Figure 2B).
2. The RMS value of the IP signal in the 10 seconds prior to the start of the episode.
3. The RMS value of the IP signal in the 10 seconds after the end of the episode.

4. The change in oxygen saturation calculated as the difference between the mean saturation in the 10 seconds prior to the start of the episode and the minimum value in the 60 seconds following the start of the episode.
5. The change in heart rate calculated as the difference between the heart rate in the 10 seconds prior to the start of the episode and the minimum value in the 60 seconds following the start of the episode.

Features 1-3 were included as it was noticed that true apnoeas often present with a marked decrease in the amplitude of the IP signal during the apnoea, which is not observed in false alarms (Figure 2A, B).

This model included input features computed before and after the apnoea to optimise it for retrospective apnoea identification. To determine whether this method could also potentially be used in prospective apnoea detection, we also adjusted the model to include only features up to 20 seconds from the start of the apnoea (including features 1 and 2, and 4 and 5 up to 20 seconds from the start as opposed to 60 seconds), see Supplementary Results.

Performance of apnoea identification

To compare the accuracy of our approach with the accuracy of the current standard, all periods where the monitor-derived respiratory rate reached 0 were viewed by two investigators and rated according to whether the investigator thought this period was a true central apnoea or a false alarm. The investigators viewed the original IP signal, the filtered IP signal, the ECG, the heart rate and the oxygen saturation for 30 seconds before and after the start of all periods where the monitor-derived respiratory rate reached 0 from all 30 infants in Data set 2.

**Supplementary Results**

*Optimising the adaptive threshold – without removal of ECG artefact*

Due to technical problems, the IP signal is occasionally recorded without the ECG signal and so cardiac-frequency noise cannot be removed from the IP signal. We assessed the performance of the adaptive threshold at different levels without the removal of ECG artefact. With the removal of ECG artefact from the signal, the optimal threshold parameters were found to be $\alpha$ = 0.4 and N=15 (see the Results). With these parameters, 9% of the manually-annotated breaths in Data set 1 were missed by the algorithm, whilst 13% of the breaths identified by the algorithm were false positives. However, a threshold of 0.7 times the standard deviation (i.e. $\alpha$ = 0.7) achieved a better balance between false positive and false negative rates without the removal of ECG artefact, 13% in each case (Supplementary Figure 4A). In the assessment of pauses in breathing, at a threshold of 0.4 times the standard deviation of the IP signal computed using the previous 15 breaths, 31% of true pauses were missed by our algorithm but only 12% of pauses identified by our algorithm were false positives. However, a threshold of 0.5 times the standard deviation computed using the previous 15 breaths, without the removal of ECG artefact, achieved a better balance between the false negative and false positive rates, at 21% and 22% respectively (Supplementary Figure 4B). Values of $\alpha$ = 0.5 and N=15 were used in the subsequent analysis (Results sections investigating the effects of morphine and ROP screening) in periods where no ECG was present (0.3% of the total ECG record in Data set 2, 0.4% of the total record in Data set 3). Values of $\alpha$ = 0.4 and N=15 were used in the results when the ECG signal was present.

*Accurate identification of apnoeas using machine learning*

The model presented in the main text included input features computed before and after the apnoea to optimise it for retrospective apnoea identification. Adjusting the model to only include features up to 20 seconds after the start of the episode still achieved an accuracy of 74% (MCC=0.43), suggesting this model could be further developed to improve prospective apnoea identification.

### Supplementary Figures

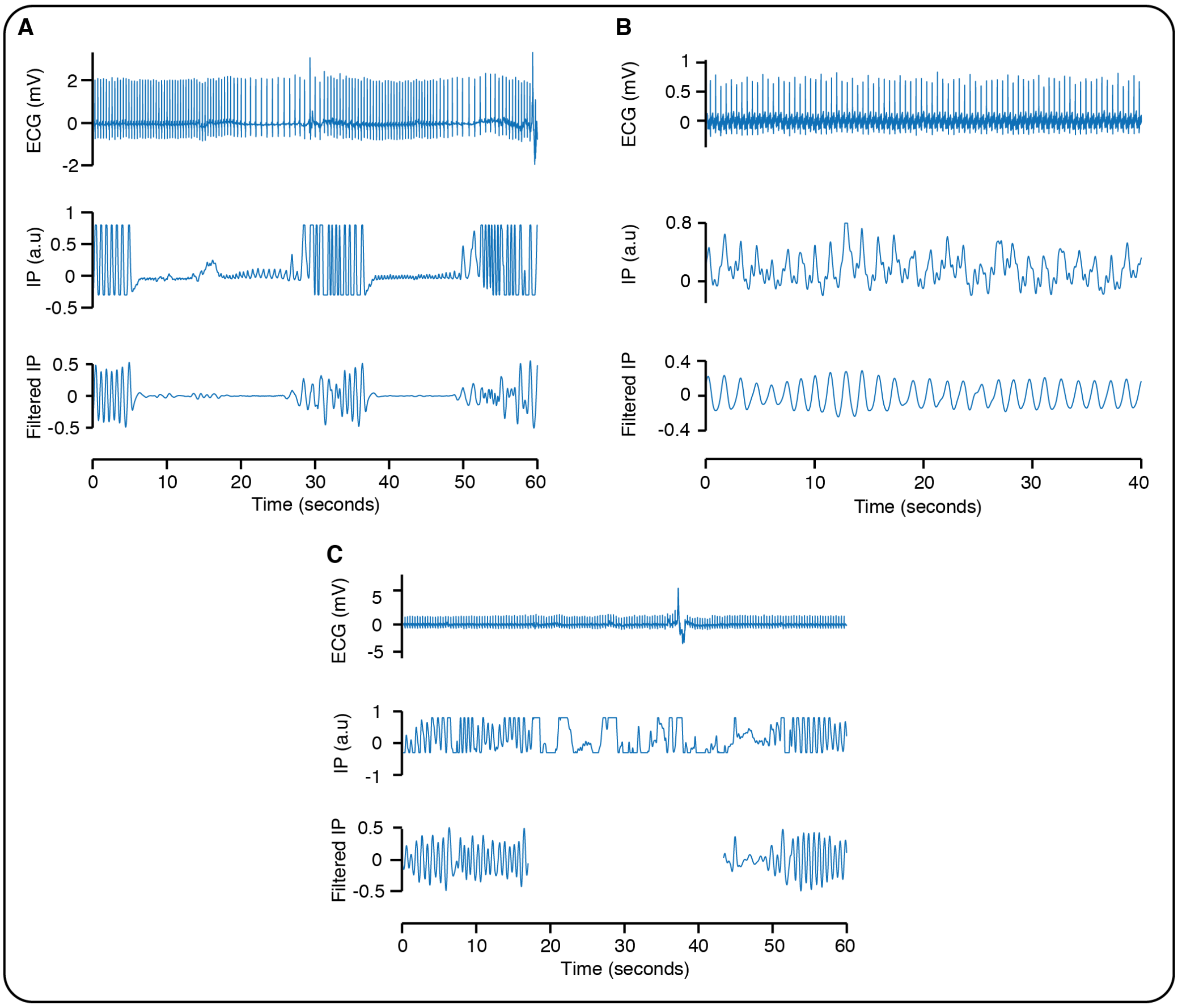


**Supplementary Figure 1: Artefact removal.** Examples of artefact removal from the IP (impedance pneumograph) signal. IP indicates the original IP signal recorded from the monitor. Filtered IP is the IP signal following removal of movement and cardiac-frequency artefacts. Time is given from the start of the epoch. (A) An example of cardiac-frequency noise which becomes particularly prominent during the period of apnoea – between approximately 15 and 25 seconds, and 40-50 seconds. (B) Ongoing cardiac-frequency noise during a period of regular breathing (the smaller amplitude oscillation on top of the higher amplitude respiratory oscillation. (C) An example of movement artefact. During periods of gross movement artefact or poor signal quality the IP signal hard limits at an upper or lower value, with the signal staying equal to this value for some period. Periods such as this were removed from the signal for analysis so they were not incorrectly identified as pauses in breathing.

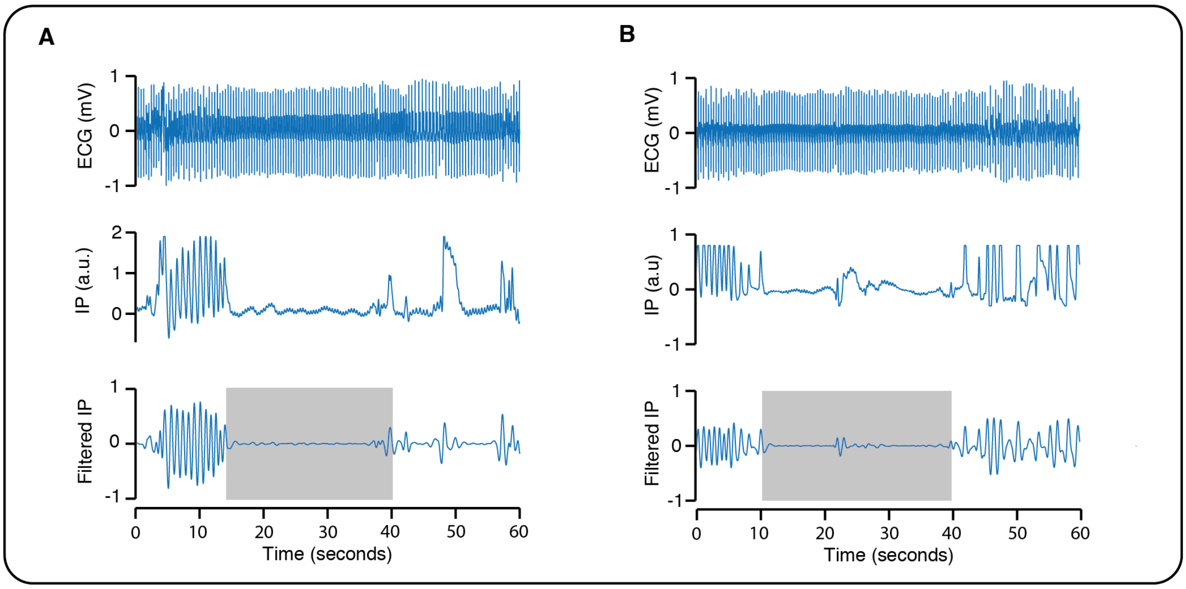


**Supplementary Figure 2: Identification of true apnoeas using machine learning.** Examples of apnoeas identified from the electrical impedance pneumograph (IP) signal. IP indicates the original IP recorded from the monitor. Filtered IP shows the IP signal following removal of cardiac-frequency noise and movement artefact. Grey shaded box indicates the period identified as an apnoea using the filtered IP. Time is given from the start of the epoch. (A) An example showing clear 'edge' effects of the IP signal at the start and end of the period identified as an apnoea, as the period identified relates to when the signal crosses a threshold (i.e. at the edges of the apnoea the amplitude of the IP signal is higher than during the rest of the apnoea). For the machine learning algorithm the first second and last second of the signal during the apnoea are removed from the analysis to avoid these edge effects. (B) An example where a single oscillation (possibly movement artefact) occurs in the middle of the apnoea. For this reason, in the machine learning analysis, the median value of the filtered IP signal, computed from the RMS values for each 1-second interval during the apnoea, is used.

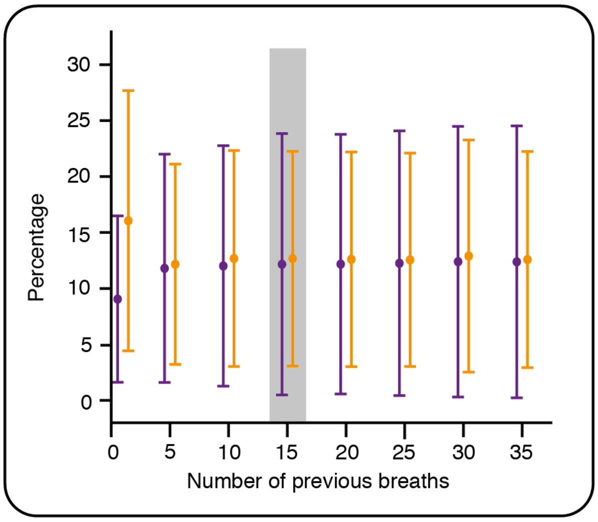


**Supplementary Figure 3: Optimising the adaptive threshold according to the number N of previous breaths.** Percentage of false positives (orange) and false negatives (purple) at different threshold values according to the number of previous breaths used to calculate the threshold. The threshold is set as 0.4 times the standard deviation of the signal from the previous N breaths. Error bars indicate mean and standard deviation. Values are jittered slightly on the x-axis so that false positive and false negative bars do not overlap. Grey shading indicates selected threshold parameters; at this threshold there was the optimal balance between the numbers of false positives and negatives.

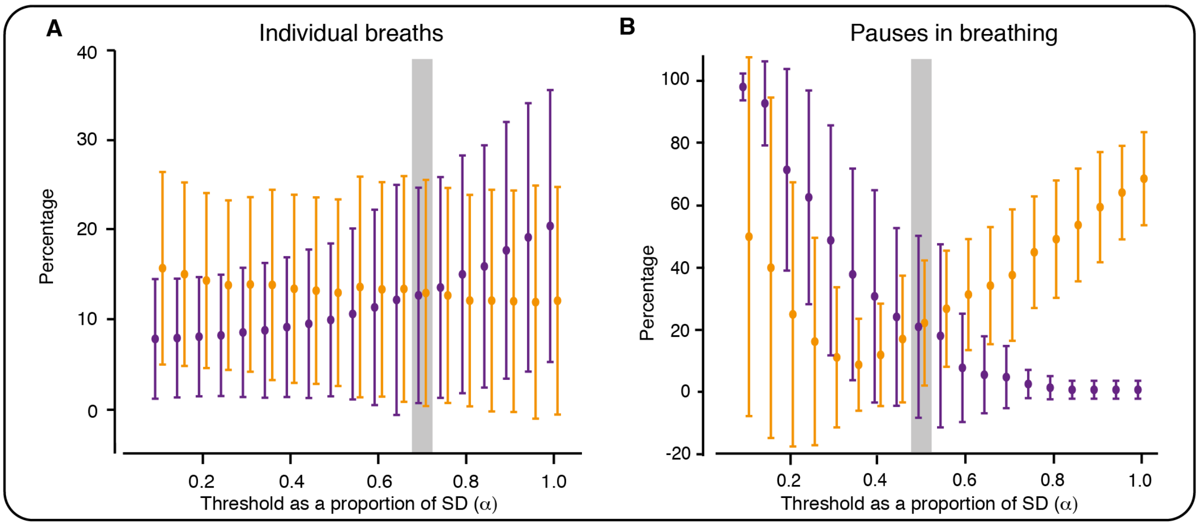


**Supplementary Figure 4: Optimising the adaptive threshold for the impedance pneumograph signal without removal of cardiac-frequency noise.** Percentage of false positives (orange) and false negatives (purple) for different values of $\alpha$ (with N=15). (A) Values calculated in relation to identification of individual breaths and compared with breaths manually annotated at the time of the recording by visual observation of the infant (Data set 1). (B) Values calculated in relation to identification of pauses in breathing compared with pauses (of length at least 5 seconds) manually annotated by two investigators from viewing the signals (Data set 2, first hour of every recording). Unlike Figure 2 cardiac-frequency noise was not removed from the signals prior to the analysis shown here. In this case, slightly higher threshold parameters achieved the optimal balance between the percentages of false positives and false negatives (see Supplementary Results). Error bars indicate mean and standard deviation. Values are jittered slightly on the x-axis so that false positive and false negative bars do not overlap. Grey shading indicates optimal value of $\alpha$ without removal of cardiac-frequency noise in both cases – the best compromise between the numbers of false positives and negatives.

### Supplementary Tables

| Procedure | Data set used |
|---|---|
| Optimising the adaptive threshold (i) by comparison with manually annotated breaths | Data set 1 |
| Optimising the adaptive threshold (ii) by comparison with pauses in breathing of at least 5 seconds | First hour of recording from 15 infants (8 allocated to receive morphine, 7 allocated to receive placebo) in Data set 2. The first hour of recording was chosen as this was not included in further analysis investigating morphine or ROP-induced changes in the IBI distribution. No drug had been given during this period. |
| Training of classification algorithm to identify true apnoeas versus false alarms | Morphine-treated infants in Data set 2 (n=15) |
| Test set for classification algorithm to identify true apnoeas versus false alarms | Placebo-treated infants in Data set 2 (n=15) |
| Testing the algorithm - evaluating respiratory depression following morphine administration | Morphine-treated infants in Data set 2 (n=15) |
| Using the algorithm - evaluating changes in IBIs following ROP screening | Placebo-treated infants in Data set 2 and Data set 3 (n=22) |

**Supplementary Table 1: Summary of where each data set was used.**

| Step | Current Study | Lee *et al.* (2012) |
|---|---|---|
| **1** | Derive RR intervals from the ECG through the application of envelopes to the ECG signal. The R-peaks are found as the points of intersect between the envelope and the ECG signal. | Derive RR intervals using the Pan-Tompkins R-peak detection algorithm ( [3], [4]). |
| **2** | Stretch or compress each RR interval such that each interval is made up of 50 steps. | Stretch or compress each RR interval such that each interval is made up of 30 steps. |
| **3** | Resample the IP in accordance with the modified RR interval series. | Resample the IP in accordance the modified RR interval series. |
| **4** | Apply a notch filter to remove the 1 Hz components from the IP, which now represent the R-peak artefact. | Apply Butterworth band-stop filters to remove R peak artefacts. |
| **5** | Apply a notch filter to remove the 2 Hz components from the IP, which represent the second harmonic of the R-peak artefact. | |
| **6** | Resample the IP to 50 Hz | Resample the IP to 60 Hz |
| **7** | High-pass filter the IP to remove <0.5 Hz noise. | High-pass filter the IP to remove <0.4 Hz noise |

**Supplementary Table 2: Filtering of the impedance pneumograph.** Differences between the computations employed in the filtering of the IP signals in this study, and those employed by Lee et al.[5].

**Supplementary References**


1 Jorge J, Villarroel M, Chaichulee S, Green G, McCormick K, Tarassenko L. Assessment of signal processing methods for measuring the respiratory rate in the neonatal intensive care unit. *IEEE J Biomed Heal Informatics* 2019; **23**: 2335–46.

2 Hartley C, Moultrie F, Hoskin A, *et al.* Analgesic efficacy and safety of morphine in the Procedural Pain in Premature Infants (Poppi) study: randomised placebo-controlled trial. *Lancet* 2018; **392**: 2595–605.

3 Monk V, Moultrie F, Hartley C, *et al.* Oral morphine analgesia for preventing pain during invasive procedures in non-ventilated premature infants in hospital: the Poppi RCT. *Effic Mech Eval* 2019; **6**. DOI:10.3310/eme06090.

4 Slater R, Hartley C, Moultrie F, *et al.* A blinded randomised placebo-controlled trial investigating the efficacy of morphine analgesia for procedural pain in infants: Trial protocol [version 2; peer review: 3 approved]. *Wellcome Open Res* 2017; **1**. DOI:10.12688/wellcomeopenres.10005.1.

5 Lee H, Rusin CG, Lake DE, *et al.* A new algorithm for detecting central apnea in neonates. *Physiol Meas* 2012; **33**: 1–17.

6 Pan J, Tompkins WJ. A Real-Time QRS Detection Algorithm. *IEEE Trans Biomed Eng* 1985; **BME**-**32**: 230–6.

7 Sedghamiz H. Complete Pan Tompkins Implementation ECG QRS detector. 2014. https://uk.mathworks.com/matlabcentral/fileexchange/45840-complete-pan-tompkins-implementation-ecg-qrs-detector.
